# Anterior Cingulate Cortex Sulcal Patterns associated with Catatonia across Schizophrenia and Mood Disorders

**DOI:** 10.64898/2026.04.20.26351285

**Authors:** Mylène Moyal, Théodor Consoloni, Alexandre Haroche, Sophie B. Sebille, Diana Belhabib, François Ramon, Adèle Henensal, Ghita Dadi, David Attali, Alice Le Berre, Clément Debacker, Marie-Odile Krebs, Catherine Oppenheim, Boris Chaumette, Anton Iftimovici, Arnaud Cachia, Marion Plaze

## Abstract

Catatonia is a severe psychomotor syndrome that occurs across psychiatric diagnoses and is increasingly conceptualized as reflecting neurodevelopmental vulnerability. The anterior cingulate cortex (ACC) plays a central role in motor initiation and cognitive–affective integration and displays substantial interindividual variability in its sulcal morphology, which is established prenatally and remains stable across life. In this MRI study, we examined whether ACC sulcal patterns represent a structural trait marker of catatonia.

We analyzed high-resolution T1-weighted images from a hospital-based cohort comprising patients with catatonia (N = 109), psychiatric patients without catatonia (N = 323), and healthy controls (N = 91). The presence of the paracingulate sulcus (PCS) in each hemisphere was determined through blinded visual inspection, and regression analyses tested associations with diagnostic group, adjusting for age, sex, scanner type, intracranial volume, and benzodiazepine and antipsychotic exposure.

Patients with catatonia exhibited a significantly reduced prevalence of the left PCS and diminished hemispheric asymmetry compared with both non-catatonic patients and healthy controls. These effects were independent of whether catatonia occurred within psychotic or mood disorders. PCS size did not differ across groups, and sulcal pattern did not correlate with catatonia severity among affected individuals.

The findings demonstrate that ACC sulcal deviations are specifically associated with catatonia across diagnostic categories, supporting a neurodevelopmental etiology and reinforcing ACC involvement in its pathophysiology. Early-determined sulcal morphology may represent a trait-level marker contributing to vulnerability for catatonia, with implications for early identification, risk stratification, and targeted intervention strategies.

## INTRODUCTION

Catatonia is a severe psychomotor syndrome that occurs across a broad range of psychiatric disorders as well as various non-psychiatric diseases (Heckers & Walther, 2023; Hirjak et al., 2024). Marked clinical heterogeneity and fluctuation of symptom expression contribute to frequent under recognition and misdiagnosis (Llesuy et al., 2018), resulting in substantial morbidity and mortality (Cornic et al., 2009; Hsu et al., 2025; Zhang et al., 2025). This is a critical issue given the availability of effective symptomatic treatments, such as lorazepam and electroconvulsive therapy (Rogers et al., 2023). Improved identification of individuals at increased risk for catatonia could therefore support earlier diagnosis, timely treatment initiation, and potentially reduce the overall disease burden.

A neurodevelopmental predisposition to catatonia has been hypothesized given its strong association with neurodevelopmental conditions (Hauptman et al., 2023) including genetic disorders (Moyal, Iftimovici, et al., 2025). Supporting this hypothesis, neuroimaging studies have reported increased quantitative gyrification in the rostral cingulate and medial orbitofrontal cortices in patients with schizophrenia and catatonia (Hirjak et al., 2019), and we recently identified qualitative deviations in orbitofrontal sulcal patterns in a similar population of patients with schizophrenia and catatonia (Moyal et al., 2024). Indeed, altered cortical sulcation and gyrification represent macroscopic markers of early neurodevelopmental deviation (Cachia et al., 2021), as cortical fold organization is largely determined in utero (White et al., 2010) and remains stable after birth (Cachia et al., 2016). In addition, we recently demonstrated electrophysiological correlates of atypical neurodevelopment in catatonia, including reduced alpha-band power and a shift in peak alpha frequency, in a transdiagnostic cohort (Moyal, Lefebvre, et al., 2025).

Converging brain imaging evidence implicates the anterior cingulate cortex (ACC) - a central hub for cognitive control, motor regulation, and emotional processing (Bush et al., 2000; Tissier et al., 2018) - as a key region in catatonia. Indeed, structural and functional ACC impairment have been repeatedly reported in catatonia, including GABAergic dysfunction during negative emotion processing and reduced connectivity with orbitofrontal regions (Northoff, 2000, 2002; Northoff et al., 1999, 2004; Richter et al., 2010), as well as structural alterations (Hirjak et al., 2019). Disruption in movement initiation and motivation in catatonia may arise from aberrant interactions between the ACC and prefrontal cortices within the broader cortico–striato–thalamo–cortical circuit (Fricchione, 2024; Fricchione & Beach, 2019).

The ACC displays considerable interindividual variability in its morphology, with two typical sulcal pattern : a “single” type - characterized by the presence of only the cingulate sulcus (CS) running along the corpus callosum - or a “double-parallel” type, which includes an additional paracingulate sulcus (PCS) running parallel to the anterior and dorsal segments of the CS (Chi et al., 1977; Ono et al., 1990), see **Figure 1**. These morphological patterns have been linked to neurodevelopmental and psychiatric disorders, as well as to cognitive impairments (Cachia et al., 2021; Subramanian et al., 2020). In particular, a reduced frequency or size of the left PCS has been reported in individuals with psychotic disorders (Fujiwara et al., 2007; J. R. Garrison et al., 2019; Lahutsina et al., 2023), individuals at ultra-high risk for psychosis (Fornito et al., 2008; Yücel et al., 2003), and individuals with bipolar disorder (Fornito et al., 2007). Moreover, patients with psychotic disorders exhibit reduced hemispheric asymmetry (Provost et al., 2003; Wu et al., 2025) which has been associated with cognitive impairments (Fornito et al., 2006; Gay et al., 2017).

**Figure 1:**
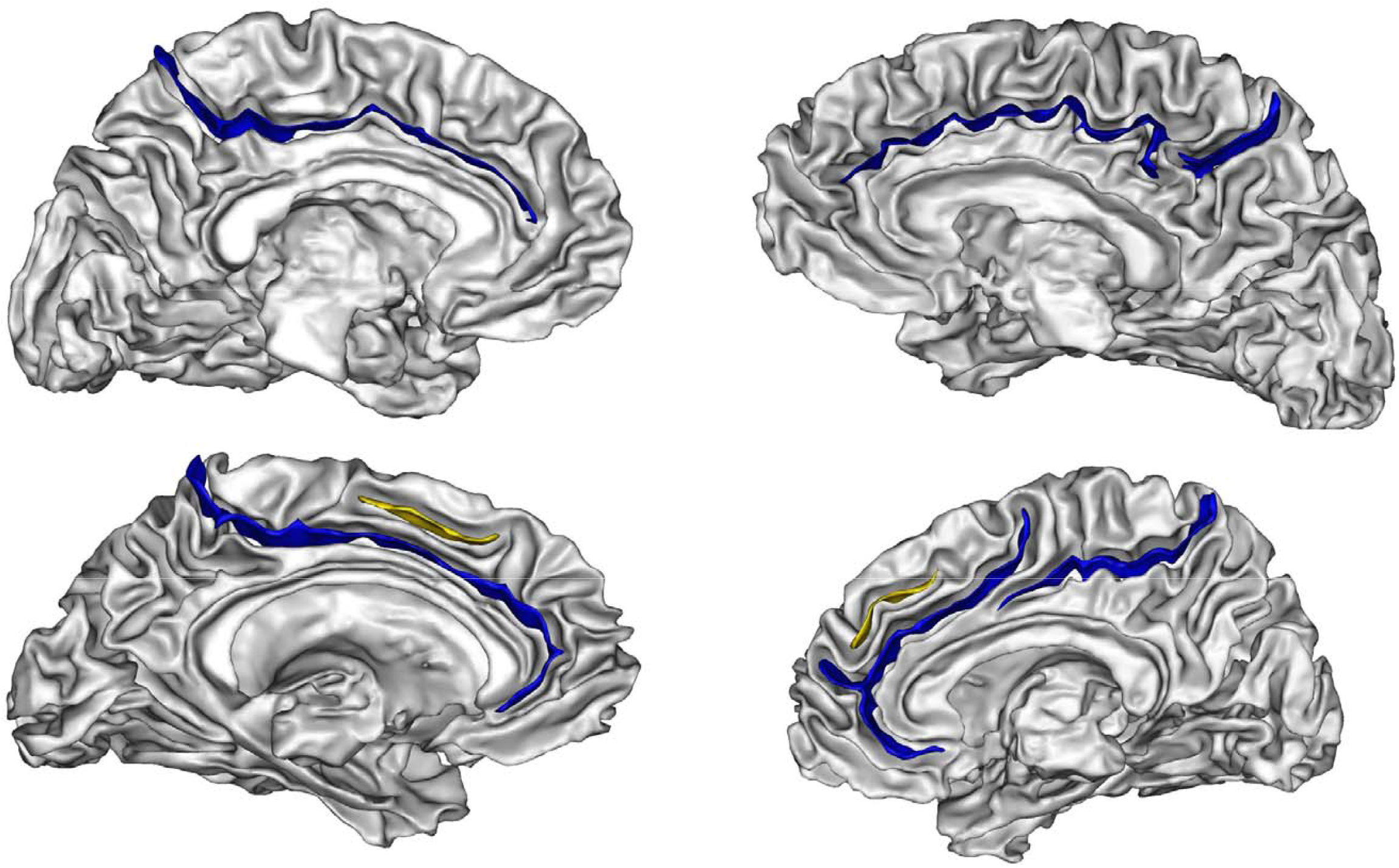
Sulcal patterns of the anterior cingulate cortex (ACC) in the left and right hemispheres. The left hemisphere is shown on the left and the right hemisphere on the right. The upper panels illustrate a ‘single’ type defined by the presence of the cingulate sulcus (CS). The lower panels illustrate the ‘double-parallel’ type, defined by the presence of an additional paracingulate sulcus (PCS) alongside the CS.

In this context, the present study aimed to further investigate the neurodevelopmental underpinnings of catatonia by examining qualitative ACC sulcal patterns in a transdiagnostic, hospital-based cohort comprising patients with catatonia (N = 109), patients without catatonia (N = 323), and healthy controls (N = 91). We tested the hypothesis that early-determined ACC sulcal patterns are altered in catatonia, independent of primary psychiatric diagnosis, thereby providing evidence for a neurodevelopmental vulnerability marker of this severe and heterogeneous syndrome.

## 1. EXPERIMENTAL PROCEDURES

### 1.1 Participants

We screened individuals diagnosed with schizophrenia or mood spectrum disorders, based on ICD-10 codes, who underwent a T1-weighted anatomical MRI between 2019 and 2024. Data were extracted from the GHU Paris Psychiatry and Neurosciences data warehouse. A subset of these patients (N = 372) had been previously included in published cohorts (Moyal et al., 2024; Moyal, Lefebvre, et al., 2025).

To identify patients with catatonia, we used automated textual research looking for the expression “catato*” or searched for ICD-10 diagnostic codes F061 “organic catatonia” or F20.2 “schizophrenic catatonia”. Each of the resulting medical records was then manually checked to confirm the diagnosis of catatonia according to DSM-5 criteria (i.e., three criteria among catalepsy, stupor, waxy flexibility, agitation, mutism, negativism, posturing, mannerisms, stereotypies, grimacing, and echo phenomena). The remaining patients were included in the non-catatonia group. The following information was extracted for every patient: age at the time of the MRI, sex, underlying diagnosis, and ongoing treatment when the MRI was undertaken, converted into equivalents of olanzapine and diazepam. This study was approved by the GHU Paris Research Ethics Committee in February 2024 (approval number 2024-CER-D-021; ‘PREDICAT’ project).

Demographic and clinical details of the study samples are reported in **Table 1**.

**Table 1.**
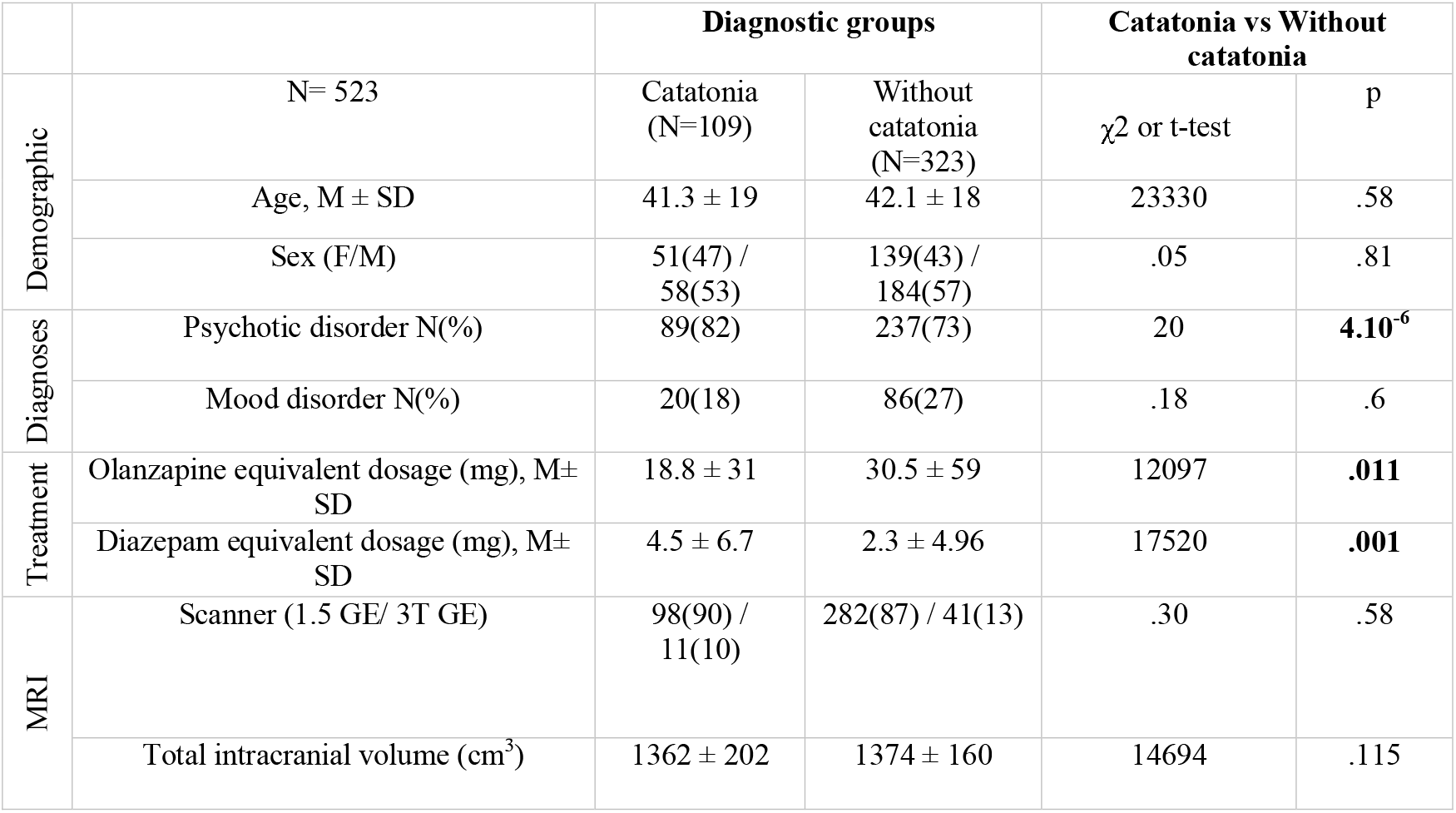
Description of the study population. Bold font indicates statistically significant difference between patients with and without catatonia. Comparisons were made using either the Z-score proportion test or the t-test, as appropriate. Results are presented as mean ± standard deviation (M ± SD) or as absolute numbers (N) with corresponding percentages.

|  |  | Diagnostic groups |  | Catatonia vs Without catatonia |  |
| --- | --- | --- | --- | --- | --- |
| | | Catatonia (N=109) | Without catatonia (N=323) | $\chi^2$ or t-test | p |
| Demographic | N= 523 |  |  |  |  |
| | Age, M ± SD | 41.3 ± 19 | 42.1 ± 18 | 23330 | .58 |
|  | Sex (F/M) | 51(47) / 58(53) | 139(43) / 184(57) | .05 | .81 |
| Diagnoses | Psychotic disorder N(%) | 89(82) | 237(73) | 20 | **4.10<sup>-6</sup>** |
|  | Mood disorder N(%) | 20(18) | 86(27) | .18 | .6 |
| Treatment | Olanzapine equivalent dosage (mg), M± SD | 18.8 ± 31 | 30.5 ± 59 | 12097 | **.011** |
| | Diazepam equivalent dosage (mg), M± SD | 4.5 ± 6.7 | 2.3 ± 4.96 | 17520 | **.001** |
| MRI | Scanner (1.5 GE/ 3T GE) | 98(90) / 11(10) | 282(87) / 41(13) | .30 | .58 |
| | Total intracranial volume (cm<sup>3</sup>) | 1362 ± 202 | 1374 ± 160 | 14694 | .115 |

Healthy controls (N=91) with (1) no psychiatric history and (2) available anatomical T1 MRI sequence, were compared to patients.

### 1.2 MRI recording

Anatomical T1-weighted MRI scans were acquired using the scanners at the Brain Imaging Facility of Sainte-Anne Hospital including 3T Canon medical (Vantage Galan 3T/XGO); and General Electric (GE) scanners (MR750, and Signa Premier), as well as a 1.5T GE scanner (see **Table 1**).

### 1.3 ACC sulcal pattern analysis

Image processing was conducted using the Morphologist toolbox in BrainVISA 4.0 (http://brainvisa.info). During automated preprocessing, T1-weighted MRI scans were skull-stripped and brain tissues were segmented. Spatial normalization was intentionally omitted to avoid biases that could result from sulcal shape distortions introduced by warping. Cortical folds were automatically delineated from the skeleton of the gray matter–cerebrospinal fluid mask; folds were defined as the crevasse bottoms of the cortical “landscape”, where height corresponds to MRI intensity. This method provides a stable and robust representation of sulcal surfaces that is not affected by variations in cortical thickness or gray/white matter contrast(Mangin et al., 2004) along with the scanner type or the acquisition protocol(Chye et al., 2017; Laval et al., 2025; Patti & Troiani, 2018). All images at each stage of processing were visually inspected, and no segmentation errors were identified. We then labelled the ACC sulcal pattern according to established inspection guidelines (J. Garrison, 2017) by annotating the CS and, when present, the PCS, including measurement of the PCS. Two independent raters (MM and TC) performed the annotations, and any discrepancies were resolved through consultation with a third rater (AC or MP). All labeling procedures were conducted blinded to participant status (i.e., catatonia, non-catatonia patients, and healthy controls). The two hemispheres have been classified independently.

The estimated total intracranial volume (eTIV) was computed in BrainVisa and incorporated as a covariate to control for potential influences of global brain volume on the observed sulcal patterns.

### 1.4 Statistical analysis

A binomial logistic regression model was fitted to examine whether the diagnostic group (“catatonia” vs. “without catatonia” vs. “healthy”) predicted PCS presence in each hemisphere. The dependent variable was the PCS pattern in the left or right hemisphere, coded as “presence” vs. “absence.”. Sex, age, scanner type (“1.5T GE”, “3T GE”, “3T Canon”), olanzapine and diazepam equivalent dose, and estimated total intracranial volume (eTIV) were included as covariates. Type III χ^2^ tests were used to account for group imbalance. Post hoc pairwise comparisons were conducted using Tukey correction to adjust for multiple testing.

In a subgroup analysis, binomial logistic regression models were again fitted with PCS presence (“presence” vs. “absence”) as the dependent variable and diagnosis subtype (“psychotic disorder with catatonia” vs. “psychotic disorder without catatonia” vs. “mood disorder with catatonia” vs. “mood disorder without catatonia” vs. “healthy”) as the independent factor, using the same covariables.

A multinomial logistic regression was then conducted to evaluate differences in PCS symmetry patterns which served as the dependent categorical variable with four mutually exclusive levels: (“asymmetry left”, with a PCS only in the left hemisphere; “asymmetry right”, with a PCS only in the right hemisphere; “bilateral presence”, with a PCS in both the left and right hemispheres; “bilateral absence”, with no PCS in the left and in the right hemispheres). The diagnostic group was the primary independent variable, and the same covariates were included.

A linear regression model was used to test whether the diagnostic group predicted PCS size, with PCS size as the dependent variable and the same covariates included.

Finally, to assess the contribution of ACC sulcal morphology to clinical expression of catatonia, linear regression models were fitted within the catatonia subgroup. Here, BFCRS score was used as the dependent variable, and sulcal patterning measures were entered as predictors, adjusting for the same covariates.

All statistical analyses were performed using the R 4.0.1 software (http://www.r-project.org) running on Rstudio with ‘nnet’, ‘multcomp’, ‘car’, ‘effects’ and ‘lsmeans’ libraries (Fox, J & Weisberg, S, 2019; Hothorn et al., 2008; Lenth, 2016; Venables, W. N & Ripley, B. D., 2002).

## 3 RESULTS

### 3.1 Demographic and clinical characteristics

A total of 523 subjects were included in the analysis (see **Table 1**).

Healthy controls (N=91) had a mean age of 29.2 ± 8 years, comprising 43 women (47%) and 39 men (43%).

Among the patient cohort, 109 individuals were diagnosed with catatonia, whereas 323 had no history of catatonia. Patients with catatonia were more likely to have a psychotic disorder compared to those without catatonia. Regarding pharmacological treatment at the time of MRI, patients with catatonia received higher doses of benzodiazepines, expressed as diazepam equivalents (4.5 mg vs. 2.3 mg; χ^2^ = 17520, p = 0.001), and lower doses of antipsychotics, expressed as olanzapine equivalents (18.8 mg vs. 30.5 mg; χ^2^ = 12097, p = 0.011). Among patients diagnosed with catatonia, the Bush–Francis Catatonia Rating Scale (BFCRS), the recommended tool for assessing catatonia severity, was available for 39 individuals, with a mean score of 20 ± 7. No difference was found regarding the scanner type and the estimated total intracranial volume.

### 3.2 Paracingulate sulcus pattern

The binomial logistic regression model revealed a significant main effect of diagnostic group (“catatonia” vs. “without catatonia” vs. “healthy”) on left-hemisphere PCS presence (χ2 = 16.65; p = .0002), but no effect for the right hemisphere (χ2 = 1.99; p = .36) after adjusting for age, sex, scanner type, total intracranial volume, and total olanzapine and diazepam equivalent doses. Post hoc pairwise comparisons (Tuckey-corrected) indicated that patients with catatonia were significantly less likely to present a left PCS compared with patients without catatonia (t-ratio = 3.8; p = .0004; Table **2** and **Figure 2**).

**Table 2.**
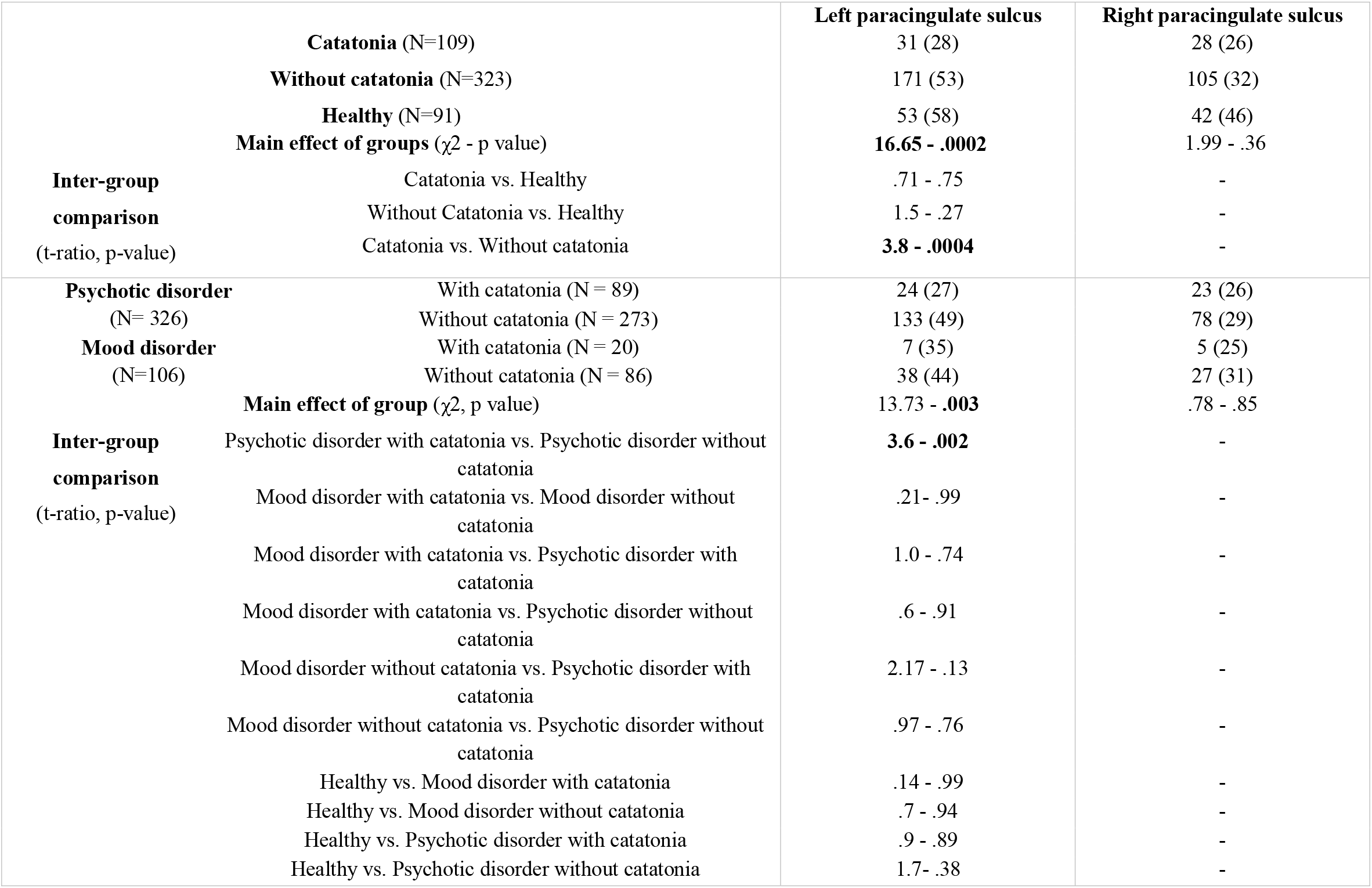
Logistic regression results for ACC sulcal pattern (paracingulate sulcus presence vs. absence) in the left and right hemispheres by group (catatonia, without catatonia, healthy) and underlying diagnosis (psychotic or mood disorder, with or without catatonia, healthy). Models were adjusted for sex, age, scanner type, olanzapine- and diazepam-equivalent doses, and total intracranial volume. Type III χ^2^ tests accounted for group imbalance; post hoc Tukey correction was applied. Significant effects are shown in bold. Results are presented as absolute numbers (N) with corresponding percentages.

**Figure 2:**
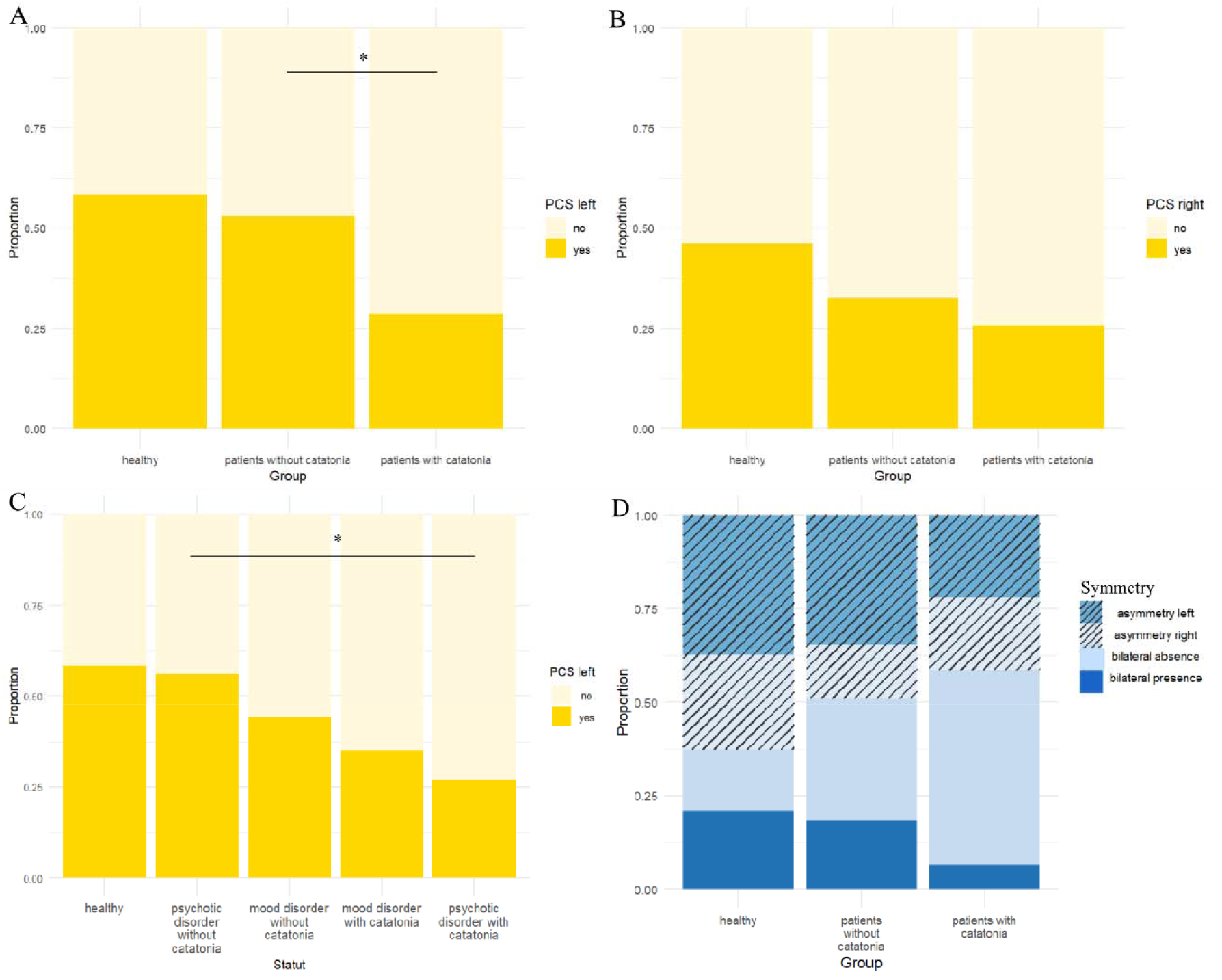
Anterior cingulate cortex sulcal pattern morphology defined by the presence or absence of a paracingulate sulcus (PCS) across diagnostic groups. (A–B) Proportion of PCS presence in the left (A) and right (B) hemispheres in healthy controls (N = 91), patients with catatonia (N = 109), and patients without catatonia (N = 323). (C) Proportion of PCS presence in the left hemisphere according to underlying diagnosis: healthy controls (N = 91), psychotic disorder with catatonia (N = 89), psychotic disorder without catatonia (N = 273), mood disorder with catatonia (N = 20), and mood disorder without catatonia (N = 86). (D) Proportion of sulcal pattern symmetry across groups: healthy controls (N = 91), patients with catatonia (N = 109), and patients without catatonia (N = 323). Asterisks indicate statistically significant differences in post hoc analyses (p < 0.05).

In the subgroup analysis by diagnosis subtype (“psychotic disorder with catatonia”, “psychotic disorder without catatonia”, “mood disorder with catatonia”, “mood disorder without catatonia”, “healthy”), a significant group effect was again observed for left PCS presence (χ^2^ = 13.73, p = .003) but not for the right PCS (χ^2^ = .78, p = .85) controlling for the same covariates. Tukey-corrected post hoc tests showed that patients with psychotic disorder and catatonia were less likely to display a left PCS than those with psychotic disorder without catatonia (t-ratio = 3.5, p = .0004; **Table 2** and **Figure 2**).

The multinomial logistic regression demonstrated a significant effect of diagnosis group on PCS symmetry pattern (χ^2^ = 13.6, p = .034), after adjustment for covariates (**Figure 2)**. However, none of the post hoc pairwise comparisons survived correction for multiple testing.

The linear regression testing PCS size found no effect of diagnosis group on PCS extent in either hemisphere (left: F = .15; p =.70; right: F = .01; p =.91).

Finally, within the subgroup of patients with catatonia, catatonia severity (BFCRS score) was not associated with PCS pattern in either hemisphere (left: F=.033; p=.85; right: F=.027; p=.87).

## DISCUSSION

In this large transdiagnostic cohort, we demonstrated that catatonia patients exhibit deviations in the anterior cingulate (ACC) morphology, reflected by a reduced prevalence of the paracingulate sulcus (PCS) in the left hemisphere and a disruption of the typical hemispheric asymmetry. These alterations were observed relative to both patients without catatonia and healthy controls, after controlling for age, sex, scanner, total intracranial volume, and total olanzapine and diazepam equivalent doses. Taken together, these findings identify an anatomical signature associated with catatonia that transcends diagnostic categories.

Cortical folding patterns are widely considered proxies of early neurodevelopmental events, as they emerge during fetal development and remain largely stable after birth (Cachia et al., 2021). The ACC sulcal deviations observed in this study therefore support the hypothesis that catatonia is linked to heightened neurodevelopmental vulnerability. Neurodevelopmental disruption in schizophrenia and mood disorders is now well established (Murray et al., 2017) and has been particularly associated with subgroups presenting more severe and treatment-resistant forms of illness (Iftimovici et al., 2024; Lefrere et al., 2025). Extending this line of evidence, our findings suggest that patients with catatonia bear an even greater neurodevelopmental burden than patients with schizophrenia or mood disorders without catatonia. This underscores the critical importance of heightened vigilance for catatonia risk in patients with a high neurodevelopmental burden (Michelini et al., 2024), especially among individuals with comorbid neurodevelopmental conditions such as autism spectrum disorder, intellectual disability, attention-deficit/hyperactivity disorder (Hauptman et al., 2023), or rare diseases (Moyal, Iftimovici, et al., 2025). For instance, individuals with 22q11.2 deletion syndrome - who exhibit altered gyrification of the anterior cingulate cortex, particularly in the context of autism (Gudbrandsen et al., 2020) - are known to be at increased risk for catatonia (Moyal, Iftimovici, et al., 2025).

Cortical folding patterns are not solely determined by biomechanical constraints related to brain size; rather, they arise from the development of cortical architecture during neurogenesis, shaped by axonal tension and regionally patterned tangential growth (Llinares-Benadero & Borrell, 2019; Tallinen et al., 2016; Van Essen, 2020). Variations in sulcal morphology may therefore reflect differences in underlying structural (Leonard et al., 2009) and functional (Fedeli et al., 2020) connectivity. As a central hub for motor control and cognitive–affective integration (Bush et al., 2000), the ACC connects prefrontal (including orbitofrontal) cortices with subcortical, striatal, and limbic structure and may act as a functional gate for the initiation and modulation of movement. In catatonia - where patients characteristically display marked impairments in initiating, sustaining, and terminating motor actions - ACC morphology, and by extension its structural and functional connectivity, may therefore be particularly consequential.

In this context, our results converge with recent observation of altered OFC sulcal patterns in patients with catatonia relative to schizophrenia without catatonia (Moyal et al., 2024), suggesting that atypical cortical folding in catatonia may extend across interconnected prefrontal regions. Deviations in ACC and/or OFC folding patterns may reflect a neurodevelopmental vulnerability within orbitofrontal–striatal– limbic networks (Fricchione & Beach, 2019; Haroche et al., 2020), potentially increasing the susceptibility of these networks to dysregulation when exposed to triggering conditions, such as acute stress, metabolic disruption or psychiatric decompensation.

The sulcal pattern of the ACC is also most likely shaped by genetic influences acting during fetal brain development (Dufournet et al., 2025), in addition to environmental factors (Mathan et al., 2024). In catatonia, a wide range of genetic variants has been reported, highlighting substantial heterogeneity in genetic susceptibility (Moyal, Iftimovici, et al., 2025). Consistently, genome-wide association studies have identified polymorphisms associated with catatonia, as well as shared genetic risk with major psychiatric disorders involving fronto-cingulate networks (Wilson et al., 2023). Taken together, these findings support the hypothesis that genetic vulnerability to catatonia may partly manifest through early neurodevelopmental alterations in cortical folding, particularly within the OFC and ACC. Such alterations could constitute a stable anatomical substrate to predispose to enduring dysfunction of cingulate–frontal circuits, ultimately contributing to the psychomotor features of catatonia.

In addition, while catatonia patients differ from both non-catatonia patients and healthy controls, patients without catatonia did not differ from healthy controls in PCS morphology, a finding that may appear counterintuitive in light of earlier schizophrenia-focused reports (Fujiwara et al., 2007; J. R. Garrison et al., 2019; Lahutsina et al., 2023). This result challenges a purely categorical diagnostic interpretation of sulcal abnormalities and instead supports a dimensional framework. In line with recent transdiagnostic evidence showing that reduced PCS asymmetry is specifically associated with hallucinations irrespective of diagnosis (Wu et al., 2025), our findings suggest that stable neurodevelopmental markers may map more closely onto symptom dimensions than to diagnostic categories - with PCS asymmetry tracking to hallucinations and left-hemisphere PCS absence more specifically associated with catatonia.

While Hirjak et al. reported hypergyrification of the rostral cingulate cortex in catatonia patients with schizophrenia (Hirjak et al., 2019), our findings indicate altered sulcogyral organization in a more caudal segment of the ACC, implicating a distinct anatomical substrate. Taken together, these observations suggest a broader deviation in ACC gyration rather than a focal abnormality confined to one subdivision. Further incorporating qualitative assessment of rostral ACC patterns is therefore warranted.

### LIMITATIONS

Several methodological considerations should be acknowledged. First, this study was retrospective, and patients did not necessarily exhibit catatonic symptoms at the time of MRI acquisition. This limitation does not undermine the interpretation of our findings, as the ACC sulcal pattern is conceptualized as a trait marker of vulnerability rather than a state-dependent biomarker of catatonia. Sulcal morphology reflects a stable neurodevelopmental characteristic and is therefore unlikely to fluctuate with transient changes in clinical status, irrespective of the timing of MRI assessment. Nevertheless, although each clinical record was carefully reviewed to confirm the presence of catatonic symptoms according to DSM-5 criteria, the retrospective nature of symptom assessment induces the possibility that some individuals with catatonia were misclassified as non-catatonic due to undocumented or unrecognized symptoms. Such misclassification would be expected to bias effects toward the null, potentially underestimating true group differences.

Second, although visual identification of the PCS followed a well-established and standardized pipeline (J. Garrison, 2017), misrecognition cannot be fully excluded. Reliance on expert visual inspection constrains scalability and limits applicability to very large datasets. Advances in self-supervised and unsupervised learning approaches offer promising avenues to automatically characterize sulcal variability at scale (Laval et al., 2025), and to enable replication of our findings in large neuroimaging databases.

## CONCLUSION

This study demonstrated deviations in ACC sulcal pattern in patients with catatonia within a large transdiagnostic hospital-based cohort. These findings underscore the contribution of neurodevelopmental vulnerability to catatonia and reinforce the involvement of the ACC in the pathophysiology and pathogenesis of this syndrome. Further work integrating genetic, developmental, and connectivity-based approaches will be critical to further elucidate the mechanisms underlying catatonia and to support earlier identification and targeted interventions.

## Data Availability

All data produced in the present study are available upon reasonable request to the authors

## ACKNOWLEDGEMENTS

Mylene Moyal has received funding from the John Bost Foundation.

This work is part of the PREDICAT project (agreement number 2024-CER-D-021, PI Mylène Moyal).

We acknowledge the contribution of Ines Bentayeb and Valentin Tran in acquiring the MRI images and organizing the data according to the BIDS standard.

